# HealthCall: Smartphone Enhancement of Brief Interventions to Improve Medication Adherence Among Patients in HIV care

**DOI:** 10.1101/2020.11.25.20235788

**Authors:** Deborah Hasin, Efrat Aharonovich, Barry Zingman, Malka Stohl, Claire Walsh, Jennifer C. Elliott, David Fink, Justin Knox, Sean Durant, Raquel Menchaca, Anjali Sharma

## Abstract

**Background:** Heavy drinking among People Living With HIV (PLWH) reduces antiretroviral adherence and worsens health outcomes. Lengthy interventions to reduce drinking and improve adherence are not feasible in most HIV primary care settings, and patients seldom follow referrals to outside treatment. Utilizing visual and video features of smartphone technology, we developed and tested HealthCall as an electronic (smartphone) means of increasing patient involvement in brief intervention to reduce drinking and improve medication adherence without making unfeasible demands on providers.

**Methods:** Alcohol-dependent patients at a large urban HIV clinic were randomized to one of three groups: (1) Motivational Interviewing (MI) plus HealthCall (n=39), (2) NIAAA Clinician’s Guide (CG) plus HealthCall (n=38), or (3) CG-only (n=37). Baseline interventions targeting drinking reduction and medication adherence were ∼25 minutes, with brief (10-15 min) booster sessions at 30 and 60 days. HealthCall involved daily use of the smartphone for 3-5 min/day, covering drinking, medication adherence, and other aspects of the prior 24 hours. Our outcome, assessed at 30 and 60 days, and 3, 6 and 12 months, was ART adherence (using unannounced phone pill-count method; possible adherence scores: 0%-100%). Analysis: generalized linear mixed models with pre-planned contrasts.

**Results:** Study retention was excellent (85%-94% across timepoints) and unrelated to treatment arm or patient characteristics. ART adherence was generally high throughout follow-up, with some decline by 12 months. Although both CG+HealthCall and MI+HealthCall evidenced benefits early in follow-up, by 6 months, ART adherence was 11% better among patients in CG+HealthCall than in CG-only (p=0.03) and 9% better than among patients in MI+HealthCall (p=0.07). Efficacy differed slightly by gender (p=.09).

**Conclusion:** HealthCall paired with CG resulted in better ART adherence than CG alone. MI+HealthCall’s early benefits diminished over time. Given the importance of ART adherence and drinking reduction among PLWH, and the low costs and time required for HealthCall, pairing HealthCall with brief interventions within HIV clinics merits widespread consideration.

## Introduction

Despite the death and disability caused by HIV over the last several decades, current antiretroviral therapy (ART) medication regimens can help patients achieve and maintain undetectable viral loads, protecting themselves from disease and preventing transmission of HIV infection to others. The finding that “Undetectable = Untransmittable” or “U = U”^1,2^ provides enormous potential to allow People Living With HIV (PLWH) to live full lives while also controlling spread of HIV infection. However, the success of U=U, Treatment As Prevention, and HIV medication in general relies on sustained high levels of patient adherence to ART regimes to maintain viral suppression^2^. Behaviors such as heavy drinking pose risks to achieving and maintaining the medication adherence that is needed for viral suppression.

Among PLWH, heavy drinking has been shown to hasten disease progression^3^, interfere with the full spectrum of engagement in HIV care^4,5^, and in particular, to interfere with HIV medication adherence^5-8^. Further, beyond unintentional nonadherence, some patients engage in intentional non-adherence due to fears of harmful effects from combining alcohol with ART^9-11^. High levels of problem drinking have been documented in PLWH treated in HIV primary care^12,13^. Among such heavily-drinking PLWH, drinking and medication non-adherence appear to be closely linked concerns that should be addressed together.

Many attempts have been made to design interventions to improve HIV medication adherence. An earlier (2013) review^14^ showed that only one of 21 adherence interventions had a strong indication of success across both adherence and clinical (i.e. CD4/viral load) outcomes. Importantly, this one study^15^ addressed alcohol and adherence together among alcohol dependent patients, suggesting potential benefit of addressing heavy drinking and poor ART adherence together. Two 2020 reviews^16,17^ both showed more promising results for ART adherence interventions than the earlier review, but also called for interventions that are cost-effective and scalable, and for more rigorously conducted trials. Many newer ART adherence interventions now incorporate mHealth technology, extending the reach of intervention, although the success of such interventions remains uncertain^18^.

Several interventions to reduce unsafe drinking in PLWH have been developed^19,20^. However, few are brief, effective, and utilize technology to extend intervention. Further, for heavily-drinking PLWH, such interventions should also address ART adherence. Therefore, utilizing the visual and video features of smartphone technology, we developed and tested “HealthCall,” an electronic (smartphone) intervention designed to enhance the efficacy of brief interventions (e.g., Motivational Interviewing (MI) and the NIAAA Clinician’s Guide [CG]) to reduce drinking and improve ART adherence among PLWH without making unfeasible demands on providers. We have already shown that HealthCall is an effective adjunct to these brief interventions in reducing heavy drinking^21^. However, the efficacy of HealthCall to improve ART medication adherence among heavy drinkers has not yet been assessed.

To test the efficacy of HealthCall to improve ART adherence in HIV-infected heavy drinkers when paired with CG or MI, we conducted a 1:1:1 randomized trial among alcohol dependent patients receiving care in an urban, HIV primary care clinic. Arm 1 received CG-only, Arm 2 received CG and HealthCall, and Arm 3 received MI and HealthCall. The primary outcome is ART adherence as indicated by a well-validated unannounced telephone pill-count method.

## Method

### Participants

Participants were recruited from and enrolled at a large urban infectious disease primary care clinic in New York City. Inclusion criteria included confirmed HIV infection, age≥18 years old, meeting DSM-IV criteria for current alcohol dependence^22^, drinking >4 drinks on at least one occasion in the past 30 days, and English- or Spanish speaking. Exclusion criteria included previous participation in a HealthCall study, active psychosis, active violent, homicidal, or suicidal thoughts, active alcohol withdrawal, gross cognitive impairment, hearing or vision impairment that precluded smartphone use, and plans to leave New York during the study period. Participants provided written informed consent. Institutional review boards at the New York State Psychiatric Institute and Montefiore Hospital, Bronx, New York approved all procedures.

### Procedures

Potentially eligible patients were informed about the study by IRB approved flyers in the clinic waiting room and/or by their providers, who referred them to meet with a study coordinator for written informed consent and assessment of eligibility. Of 180 individuals screened in-person with the Psychiatric Research Interview for Substance and Mental Disorders for DSM-IV (PRISM-IV-Computerized Version)^23^, 114 met eligibility, completed baseline assessments, and were randomized (see Hasin et al., 2020 MedRxiv for further detail on study enrollment)^21^. In a parallel three-arm 1:1:1 randomized design, participants were assigned to one of three conditions: CG-only (n=37), CG +HealthCall (n=38), or MI+HealthCall (n=39). Randomization was stratified on drug use severity, depression, and unstable housing using urn randomization^24^. All baseline assessments were completed prior to randomization. After each study visit, participants were compensated via ClinCards for their completed assessments. All participants also received a study smartphone or the equivalent value in gift cards at their 60-day visit, the end of treatment.

### Interventions

Study counselors, part of the regular clinic staff, delivered the interventions after receiving training in their delivery. A clinic health educator delivered the MI. An RN or a physician-fellow delivered CG. The baseline MI and CG sessions took equivalent amounts of time (∼25 minutes). While delivering CG in the CG-only and CG+HealthCall arms, the counselors were blinded to participants’ assignment to HealthCall; about 20 minutes into the CG session, a study coordinator texted the counselor to inform them if the patient was randomly assigned to receive HealthCall. If assigned, the counselor then introduced the HealthCall app to the patient. This two-step randomization process kept counselors blinded to the HealthCall assignment until after the CG was delivered to avoid knowledge of the assignment influencing the way counselors delivered CG. Intervention sessions were audiotaped and reviewed for quality assurance. Participants returned at 30 and 60 days after the baseline intervention for assessments and a brief booster session (10-15 minutes). Total duration of the treatment period was 60 days across all three arms. Further face-to-face follow-up assessments were conducted at 3, 6, and 12 months after baseline.

### Baseline Clinician’s Guide (CG)

The evidence-based CG^25-27^ was designed and recommended by the National Institute on Alcoholism and Alcohol Abuse (NIAAA) for use by providers such as medical personnel^28^, including those in HIV care settings.^29^ Consisting of a flow-chart to guide a set of recommended drinking-reduction techniques^30^, CG requires little training and staff time.^31^ The CG was adapted for this study to address slightly lower drinking reduction goals due to HIV infection, and also to address ART adherence. The ART adherence components were structured based on the IAPAC Guidelines Regimen Information Program (GRIP), an evidence-based approach^32^. This approach uses a screener for ART adherence in the prior 30 days^33,34^, and step-by-step guidance based on common^35^, evidence-based^36^ tools and recommendations of clinic resources^35,36^. Because the CG and GRIP approaches were so similar, the GRIP was readily integrated into the Clinician’s Guide. Using this adapted version of the CG, patient ART adherence was queried, associated consequences and barriers discussed, and patients were encouraged to track and work on improving their ART adherence.

### Baseline Motivational interviewing (MI)

The brief MI session followed standard MI techniques^37^, e.g., exploring ambivalence about drinking reduction, health consequences of drinking and medication nonadherence, and setting a drinking goal for the next 30 days. Patients were also queried about ART adherence and barriers to improvement, and MI techniques were used to encourage improving ART adherence.

### HealthCall

Patients randomized to HealthCall arms received an Android smartphone on which the HealthCall application and a pre-paid calling and data plan were installed. Study counselors introduced patients to HealthCall by explaining its purpose, instructed them in its use and that this took ∼3-5 min each day, and asked them to try it for the first time on-site. Patients were instructed to use HealthCall daily for the next 30 days (and at 30-day booster, were asked to use HealthCall for another 30 days). HealthCall is proposed to work through two features: self-monitoring and personalized feedback. ***Self-Monitoring***: The HealthCall script includes self-monitoring questions in English or Spanish (patient’s preference) about the previous 24 hours^38^. The interactive queries asked about ‘yesterday’ (morning, afternoon, evening) to ensure consistent reporting periods regardless of the time patients accessed HealthCall. HealthCall queried several aspects of health, including drinking quantities, reasons for drinking/abstaining, and whether they took their ART medication that day. ***Personalized feedback*:** Patient’s daily HealthCall data were automatically transmitted to a secure server, compiled, and used to produce personalized graphs representing the number of standard drinks patients drank each reported day relative to their drinking-reduction goal, as well as the days missed medication (which can be aligned with drinking graph to determine drinking/adherence links). The 30 and 60-day booster sessions included a review and discussion of these graphs.

### Brief booster sessions at 30- and 60-days

Following baseline CG or MI, patients in CG+HealthCall and MI+HealthCall used the HealthCall app daily. At 30- and 60-day visits, patients in all three arms met with the clinic staff member again for a booster session to review drinking and medication adherence over the prior 30 days, and reinforce any improvement and continued efforts. In the CG-only arm, discussion was based on patients’ recall. In the CG+HealthCall and MI+HealthCall arms, discussion was based on a graphed report of daily drinking and ART adherence generated from the HealthCall app data. At the end of the 30-day booster visit, any revised drinking goals were entered into the app by the counselor. Patients were then asked to continue using HealthCall for another 30 days. At the end of the 60-day session (end of treatment), options for continued alcohol care were discussed, and referrals given to patients who wished to receive them.

## Measures

At baseline, 30 and 60 days, prior to receiving study interventions, participants completed a 60-minute computer-assisted personal interview (CAPI) that was administered by a trained study assessor who was not involved in administering the interventions. The CAPI was also completed at 3, 6 and 12 months after baseline. The CAPI covered demographic characteristics, DSM-IV substance and psychiatric disorders, and other medical and HIV-related health variables. To identify any patients with detoxification needs alcohol withdrawal was assessed at baseline, 30- and 60-days visits with the Clinical Institute Withdrawal Assessment of Alcohol revised (CIWA-Ar)^39^.

All participants taking ART medication were assessed for medication adherence eight times after the baseline. This assessment consisted of the unannounced pill count phone survey method^40-42^, administered by a trained study interviewer. During these pill count surveys, patients counted the number of pills they has of each HIV medication. The calls were completed when patients were at home, and not scheduled in advance. This established method for assessing ART medication adherence has good reliability and validity^40-42^, predicts viral suppression^40^, and avoids the overestimates of adherence^43^ from using ACTG-like measures.

To prepare for these pillcount assessments at baseline, study staff trained patients on how to count their ART medication using a pill count tray we provided, and established the patient’s most convenient time of day for the subsequent pillcount calls. The first of these calls occurred a few days after the baseline visit. The next three calls occurred around the 30-day, 60-day and 3-month points after baseline, and were 28-35 days apart. The next four calls occurred around 5 months, 6 months, 11 months, and 12 months after the baseline visit. If the participant was not available/home at the time of the initial call, the interviewer called again at additional times. For the purpose of the current study, we used 30-day, 60-day, 3 month, 6 month, and 12 month data, to parallel the timeframes analyzed in our primary drinking analysis^21^.

### Outcomes

ART adherence is calculated using a formula accounting for all pills scheduled compared to pills on hand since last count. Adherence (range, 0.0-1.0) is a function of prescribed doses relative to pill count. Given the 2016 WHO “Treat All” recommendations^44,45^, adherence is recorded as 0.0 for those not taking ART due to lack of a prescription. Adherence levels are defined as high (81%-100%), medium (41%-80%), and low (0-40%). These cutoffs are consistent with scientific guidelines, as those with <40% adherence are most at risk for drug resistance^46^, whereas ∼82% adherence has been deemed necessary to attain viral suppression in most cases^47^.

### Statistical analysis

Baseline demographic characteristics were examined descriptively among the whole sample and by treatment group. Differences in characteristics by treatment group were tested using non-parametric Wilcoxon tests for continuous variables and Chi-square tests for categorical variables.

Change in medication adherence was analyzed using unannounced pill count data from assessments during treatment (30 and 60 days) and from the follow-up assessments (3, 6, and 12 months). A generalized linear mixed model (GLMM) was used to model the effects of time, treatment, and the interaction of time x treatment on adherence using SAS PROC GLIMMIX^48^. Models used a normal distribution to model the outcomes. Pre-planned contrasts were used to compare treatment groups (CG+HealthCall vs. CG-only; MI+HealthCall vs. CG-only; MI+HealthCall vs. CG+HealthCall). Tests were two-sided with p<0.05 indicating significance, also noting trends toward significance at p<0.10. Participants were all analyzed in their originally assigned treatment group. Estimated mean differences were used to represent effect sizes. A potential interaction with gender was assessed by including the three-way interaction term of time x treatment x gender and associations within each gender were then explored separately.

## Results

### Sample characteristics

As described elsewhere^21^, of the 114 enrolled patients, 58% were male, 75% were African American, 28% were Hispanic, and 62% had less than a high school education. The mean age was 47.5 years (SD = 10 years) and the mean number of years of HIV infection was 18.6 (SD =7.6). Thirty-seven patients were randomized to CG-Only, thirty-eight to CG+HealthCall, and thirty-nine to MI+HealthCall. One patient randomized to CG-Only did not keep an appointment for the baseline intervention and thus was not exposed to any protocol intervention; this participant was not included in the analyses. None of the baseline demographic characteristics differed significantly by treatment condition.

### Study retention

Overall, study retention was high across the treatment conditions. Of the 114 participants, 98 (86%) completed the study (n=31 in CG-Only, n=36 in CG+HealthCall, and n=31 in MI+HealthCall). Retention did not differ significantly by treatment condition or any baseline characteristic.

Participants reported high utilization of HealthCall. Among patients in MI+HealthCall, the median HealthCall call rate was 89.17%. Among participants in CG+HealthCall, the median HealthCall call rate was 87.50%.

The overall response rate for completion of all possible pill count surveys was 97.6% (799 out of 819 that could have been completed). The response rate ranged from 100% just after baseline to 97% at 12 months.

### ART adherence

More than 70% of the patients reported high levels of adherence (81-100%) at 30 day, 60 day, 3 month, and 6 month follow-ups, with fewer (63.2%) at this level at the 12 month follow-up (Table 1). Few participants (4-6%) reported low adherence (0%-40%) at most timepoints, with the exception of the 6 month follow-up, when 12.4% reported low adherence.

**Table 1.** Adherence levels at each time point: Unannounced pill count data.

| Mean Adherence<br>score level<br>(% of ART they took)<sup>a</sup> | % of patients in each category |  |  |  |  |
| --- | --- | --- | --- | --- | --- |
|  | 30-day<br>N=105 | 60-day<br>N=100 | 3-month<br>N=101 | 6-month<br>N=97 | 12-month<br>N=95 |
| **Low**<br>0% – 40% | 5.7 | 4.0 | 5.9 | 12.4 | 5.3 |
| **Medium**<br>41% - 80% | 23.8 | 21.0 | 21.8 | 17.5 | 31.6 |
| **High**<br>81% – 100% | 70.5 | 75.0 | 72.3 | 70.1 | 63.2 |
| **Range** | 28% - 100% | 13% - 100% | 29% - 100% | 7% - 100% | 21% -<br>100% |
| <sup>a</sup> Perfect adherence = 100% |  |  |  |  |  |

Overall, ART adherence declined slightly over time (see Figure 1a). Groups evidenced no difference in adherence at the 30-day follow-up (Table 2). At 60 days, CG+HealthCall evidenced a 10% greater adherence than those in CG-only, p<0.05, and MI+HealthCall evidenced a marginally (9%) greater adherence than the CG-only group, p=0.07. At 3 months, there was a marginal benefit of CG+HealthCall over CG-only (9%; p=0.09). However, by 6 months, ART adherence was better in CG+HealthCall than it was in both CG-only (11%; p=0.03) and in MI+HealthCall (9%; p=0.07). No group differences remained at 12 months.

**Table 2.** Differences in ART adherence (measured by unannounced pill count) by treatment group.

|  |  | Contrasts |  |  |  |  |  |
| --- | --- | --- | --- | --- | --- | --- | --- |
|  |  | CG+HealthCall<br>vs. CG Only |  | MI+HealthCall<br>vs. CG Only |  | MI+HealthCall<br>vs. CG+HealthCall |  |
|  |  | Estimated<br>Mean<br>Difference | p-value | Estimated<br>Mean<br>Difference | p-value | Estimated<br>Mean<br>Difference | p-value |
| **% Medication Adherence** |  |  |  |  |  |  |  |
|  | 30 days | 0.05 | 0.30 | -0.010 | 0.85 | -0.06 | 0.22 |
|  | 60 days | 0.10 | 0.048 | 0.09 | 0.065 | -0.007 | 0.89 |
|  | 3 months | 0.09 | 0.09 | 0.06 | 0.27 | -0.03 | 0.54 |
|  | 6 months | 0.11 | 0.03 | 0.02 | 0.72 | -0.09 | 0.07 |
|  | 12 months | 0.06 | 0.23 | 0.06 | 0.26 | -0.004 | 0.94 |

Note. Model was run as a linear mixed model .

**Figure 1a.**
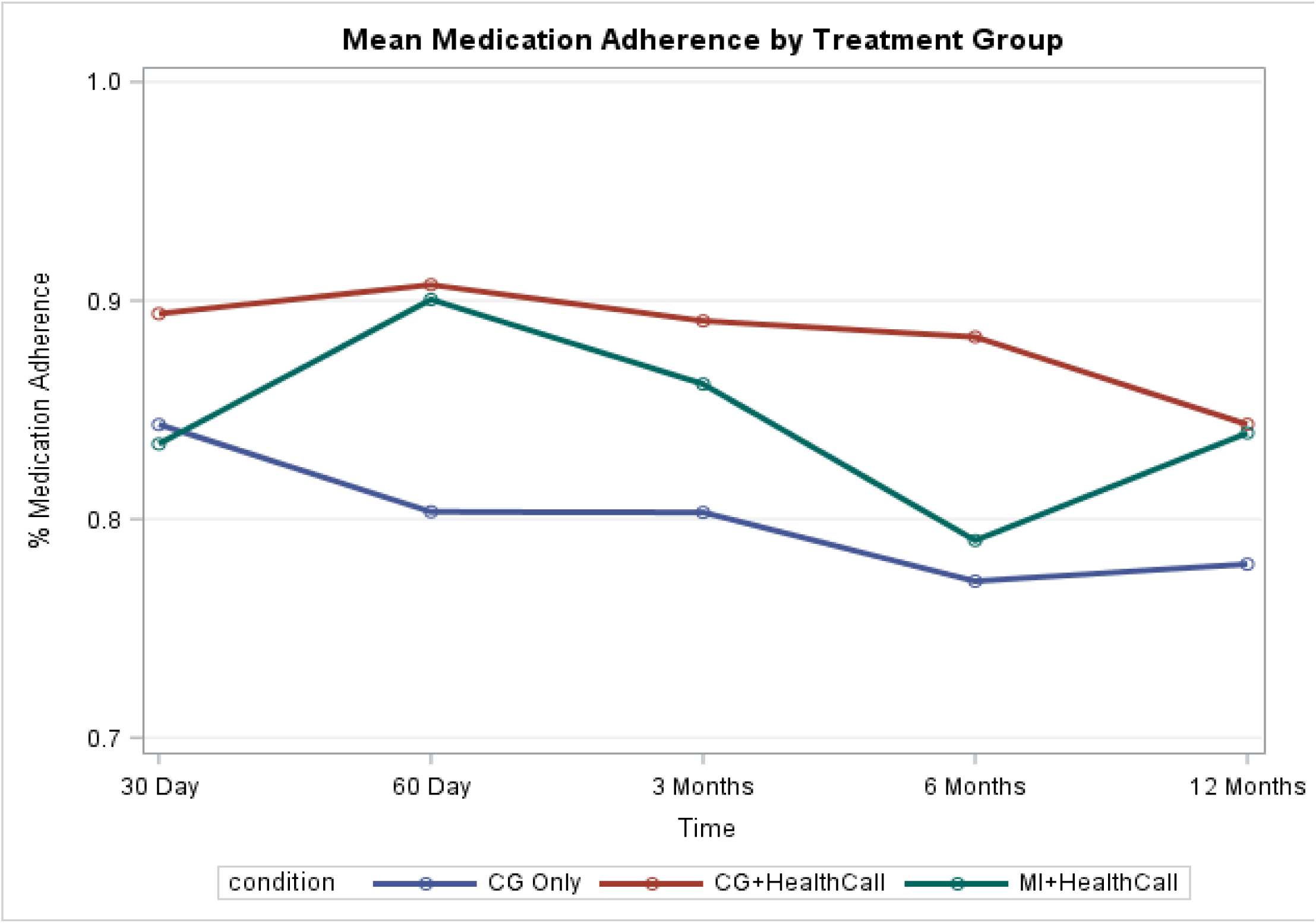
Mean medication adherence by treatment group.

**Figure 1b.**
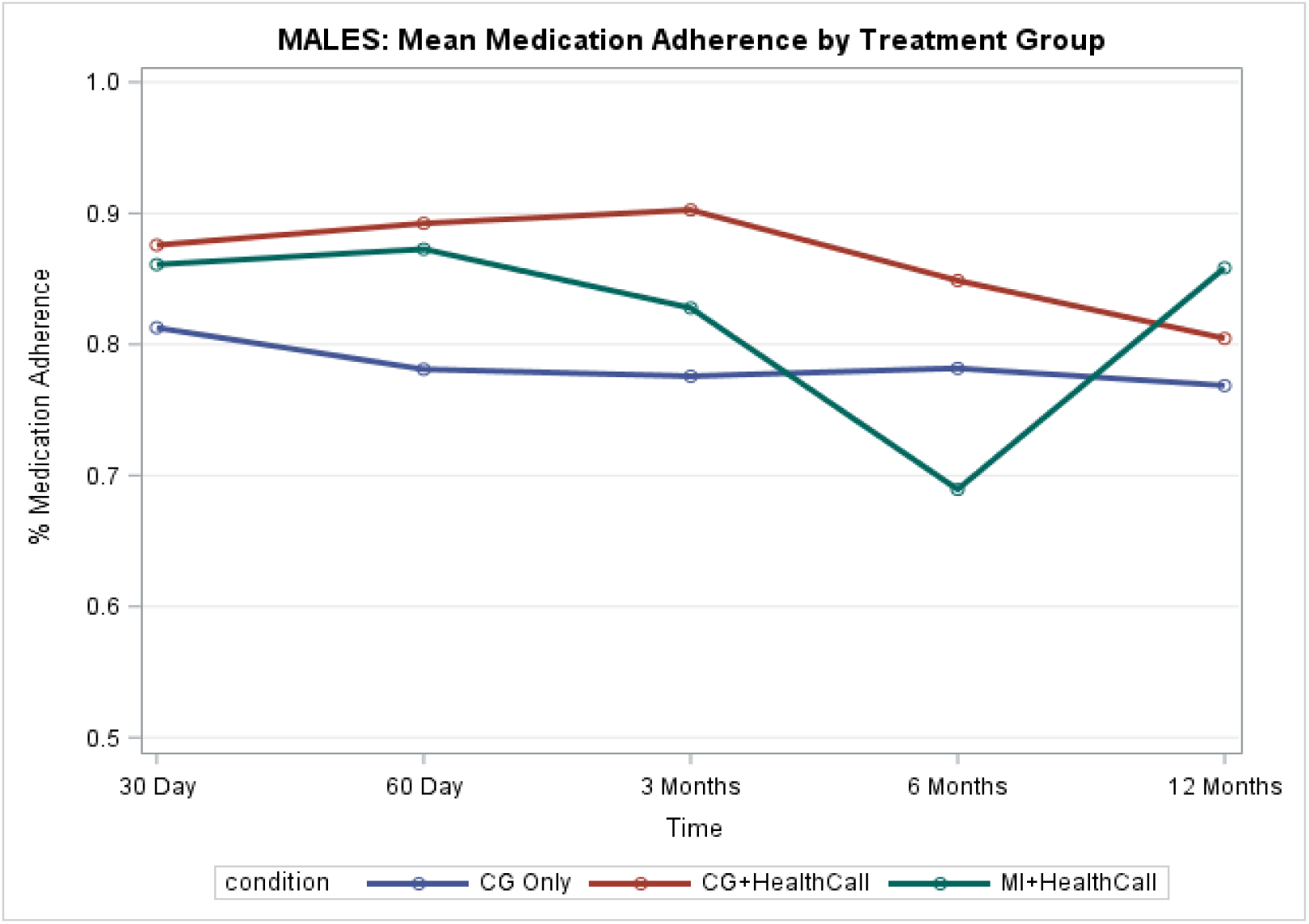
Mean medication adherence by treatment group among males only.

**Figure 1c.**
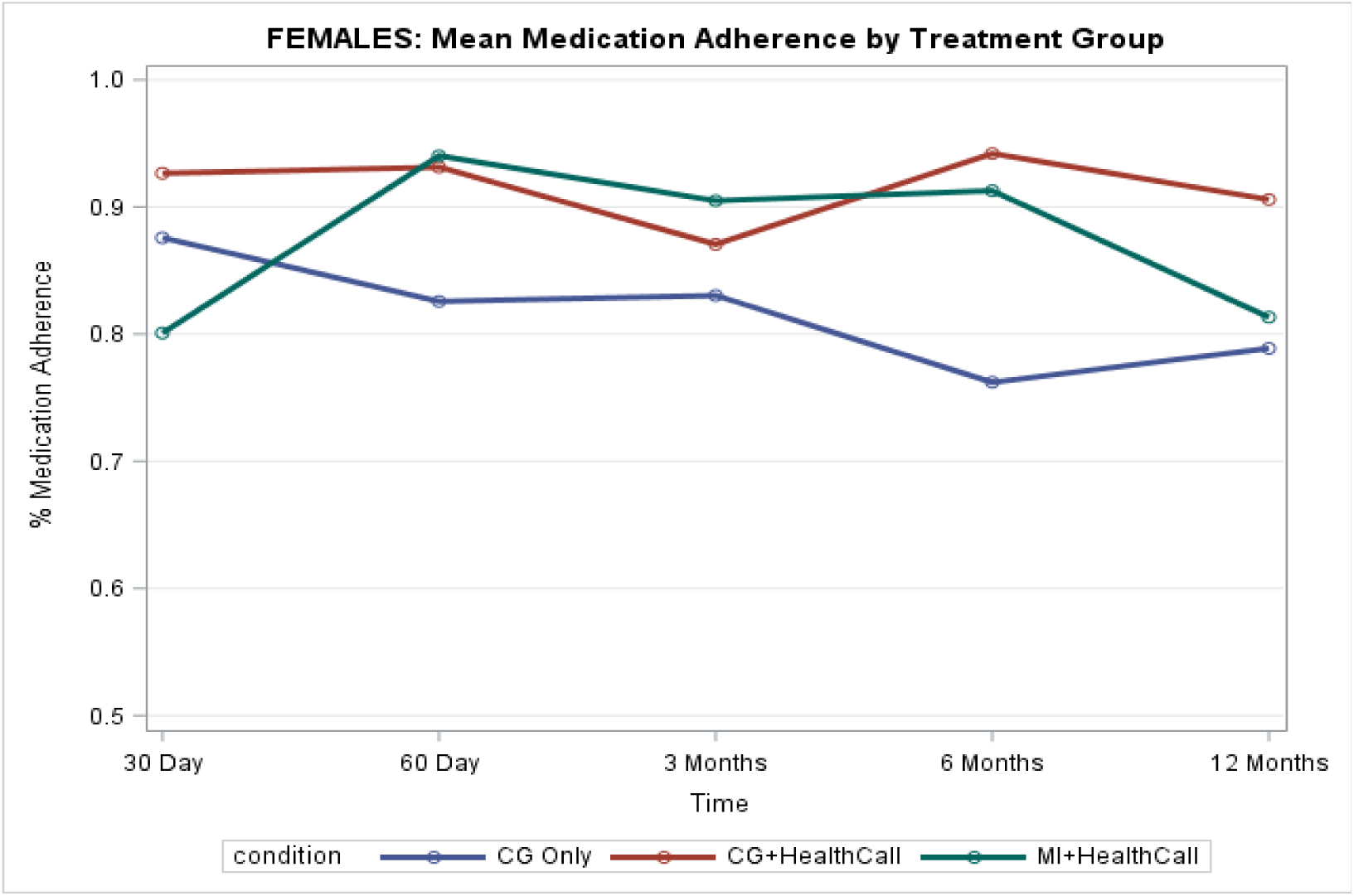
Mean medication adherence by treatment group among females only.

**Table 3.**
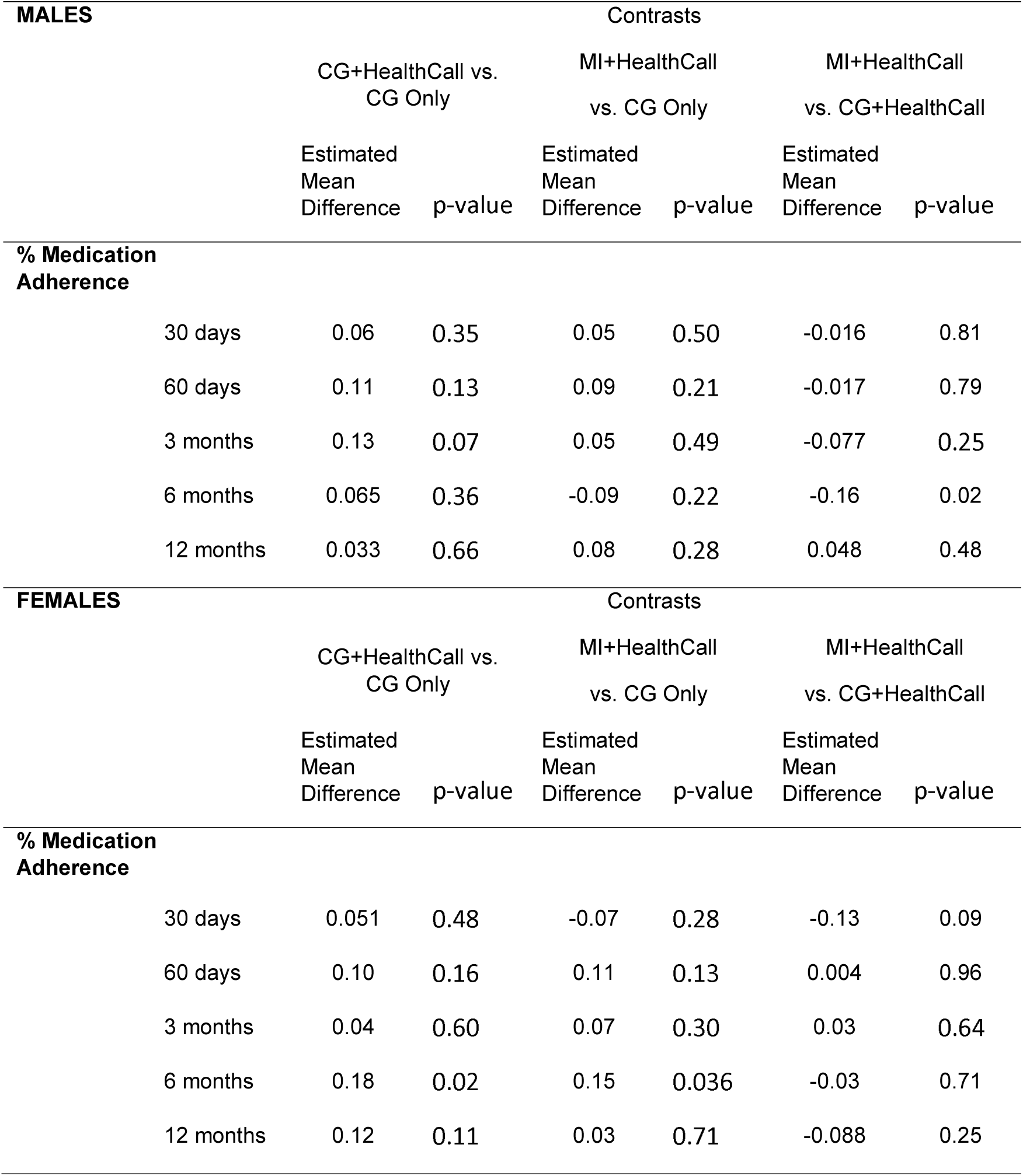
Differences in ART adherence (measured by unannounced pill count) by treatment group and by gender.

### Interaction by gender

There was a marginal interaction of condition by time by gender on ART adherence, *F*(8, 368) = 1.72, p=0.09. Males and females were thus examined separately. ***Males***. Among men, at 3 months, those in CG+HealthCall had marginally better adherence than in those in CG-Only (13%; p=0.07). At 6 months, those in CG+HealthCall reported better adherence than those in MI+HealthCall (16%; p=0.02). ***Females***. Among women, adherence was marginally better among participants in CG+HealthCall than in MI+HealthCall at 30 days (13%; p=0.09). Although groups were similar at 60 days and 3 months, by 6 months, participants in both CG+HealthCall (18%; p=0.02) and MI+HealthCall (15%; p=0.04) outperformed CG-only.

## Discussion

This study indicates that brief intervention (specifically the NIAAA Clinician’s Guide [CG]) can be supplemented with the HealthCall smartphone app in order to improve adherence to ART medication. For men, CG+HealthCall was superior; for women, both interventions paired with HealthCall provided benefit compared to CG-only 6 months later. The use of a smartphone app can thus extend the reach of a brief drinking and adherence intervention, providing continued care without further demands on providers’ time, which can be costly and in high demand.

Although many interventions have been developed to address drinking^19,20^ and ART adherence^14,16-18^ among PLWH, many require substantial provider time or show limited efficacy. Given the well-documented relationship between heavy drinking and poor ART adherence^5-8^, our intervention addresses these two concerns together among alcohol dependent PLWH. The current intervention utilizes smartphone technology to provide an extended intervention without taxing providers’ time, thus providing a feasible intervention method in HIV primary care.

Study limitations are noted. Recruitment took place at one urban clinic in the northeast, with primarily Black, male participants, limiting generalizability to other groups. The population was alcohol dependent, and the intervention had a substantial focus on drinking reduction, so the study does not provide information on whether HealthCall is a useful tool for improving medication adherence among non-drinkers; further research would be required to determine this. Also, we did not include an MI-only arm in the current study because reviewers of the grant proposal for this study felt that its inferiority to MI+HealthCall using an earlier, non-smartphone version of HealthCall had already been demonstrated^49^. However, this clinical trial as well as the HealthCall intervention have several important strengths. Our study design tested HealthCall as an adjunct to two brief interventions, allowing us to assess comparability of a more disseminable/scalable intervention (CG) than one that requires more training, supervision, and clinical skill (MI). Our study evidenced high retention through follow-up, with high participation in pill count phone surveys. The unannounced pill count phone survey method we used has demonstrated reliability and validity, and minimizes overestimation in comparison with memory-based self-report. The HealthCall app itself also has many strengths, as it improves efficacy and requires minimal provider involvement, harnessing technology to extend the reach of brief provider contact.

In sum, this research provides evidence that the HealthCall administered via smartphone can extend the reach of brief intervention in decreasing drinking^21^ and increasing medication adherence among alcohol-dependent PLWH, providing a valuable tool that can enhance several aspects of health together among alcohol dependent PLWH. Technology has become increasingly useful and well-recognized in managing health. In particular, the importance of telehealth has become acutely clear since the start of the COVID-19 pandemic, as it can supplement HIV and other care when on-site care is unavailable, infeasible, or risky^50^. Telehealth has grown in popularity during COVID-19, and its demonstrated utility and ease is likely to lead to continued widespread use. HealthCall similarly uses technology to enhance patient care, thus serving as a useful add-on to in-person (or perhaps even online) counseling to improve health outcomes among alcohol dependent PLWH.

## Data Availability

Patient data is confidential and protected. Please contact with questions.

